# Dose-response effect of dietary nitrate on muscle contractility and blood pressure in older subjects

**DOI:** 10.1101/2020.05.29.20117218

**Authors:** Edgar J. Gallardo, Derrick A. Gray, Richard L. Hoffman, Brandon A. Yates, Ranjani N. Moorthi, Andrew R. Coggan

## Abstract

**Purpose:** We have recently demonstrated that dietary nitrate, a source of nitric oxide via the enterosalivary pathway, can improve muscle contractile function in healthy older men and women. Nitrate ingestion has also been shown to reduce blood pressure in some, but not all, studies of older individuals. However, the optimal dose for eliciting these beneficial effects is unknown.

**Methods:** A randomized, double-blind, placebo-controlled crossover study was performed to determine the effects of ingesting 3.3 mL/kg of concentrated beetroot juice (BRJ) containing 0, 200, or 400 µmol/kg of nitrate in nine healthy older subjects (one man, eight women; mean age 70±1 y). Maximal knee extensor power (Pmax) and speed (Vmax) were measured 2 h after BRJ ingestion using isokinetic dynamometry. Blood pressure was monitored periodically throughout each study.

**Results:** Pmax (in W/kg) was higher (P< 0.05) after the low dose (3.9±0.4) compared to the placebo (3.7±0.4) or high dose (3.7±0.4). Vmax (in rad/s) also tended higher (P = 0.08) in the low (11.9±0.7) compared to the placebo (10.8±0.8) or high dose (11.2±0.8) trials. Eight out of nine subjects achieved a higher Pmax and Vmax after the lower vs. the higher dose. These dose-related changes in muscle contractility paralleled changes in nitric oxide (NO) bioavailability, as reflected by changes in breath NO and plasma 3-nitrotyrosine levels. No significant changes were found in systolic, diastolic, or mean arterial pressure.

**Conclusions:** Varying doses of dietary nitrate have differential effects on muscle function and blood pressure in older individuals. A moderate dose of nitrate increases muscle speed and power, but these improvements are lost at a higher dose. Blood pressure, on the other hand, is not reduced even with a higher dose.

---

Roughly ⅓ of individuals over age 65 y suffer from some form of ambulatory or self-care disability (1). In the United States alone, this amounts to some 15 million individuals at present, a number is likely to at least double over the next three decades (1). Similar trends are evident world-wide (1). The primary cause of disability in older persons is a decline in muscle contractile function, with serious difficulty walking or climbing stairs being the most prevalent problem at all ages (1). These functional limitations are highly predictive of dependence, institutionalization, and mortality in the elderly (2,3). Thus, it is clear that age-related changes in muscle function can have an enormous impact not only on a given individual’s quality and length of life but also on the societal costs of caring for an aging population.

Numerous factors contribute to the decline in muscle contractile function with aging. These include a reduction in overall muscle mass, as a result of a decrease in both muscle fiber number and size, as well as alterations in the neural control of muscle (4,5). These contribute to a decrease in strength, i.e., a reduction in the maximal force-generating capacity of muscle. However, the maximal speed of muscle shortening also decreases with aging (6–9), leading to an even more dramatic age-related decline in maximal muscle power, i.e., the product of force and speed (7,8). Moreover, decreases in muscle power with aging are a better predictor of movement disabilities than decreases in strength (9,10). Improving muscle speed and power should therefore logically be the primary focus of any intervention intended to combat muscle dysfunction in the elderly.

We have previously reported that dietary nitrate (NO_3_^−^), a source of nitric oxide (NO) via the reduction of NO_3_^−^ to nitrite (NO_2_^−^) and hence to NO via the action of bacterial NO_3_^−^reductases in the mouth and/or mammalian NO_3_^−^ reductases (e.g., xanthine oxidase (XOR)) in the tissues (10), can significantly enhance muscle speed and power in various subject populations, i.e., healthy young and middle-aged individuals (11,12), athletes (13), patients with heart failure (14). Although the exact mechanisms responsible for this effect are still uncertain, they appear to involve NO-mediated changes in muscle calcium (Ca^2+^) release and/or sensitivity (15). More recently, we have demonstrated that acute NO_3_^−^ ingestion also significantly improves muscle speed and power in healthy elderly subjects (16). These changes were large enough to theoretically offset the reductions resulting from several decades of aging. It is therefore clear that dietary NO_3_^−^ supplementation holds enormous potential as a means of mitigating muscle dysfunction, and hence disability, in older individuals. The optimal dose of NO_3_^−^ for eliciting improvements in muscle contractility in elderly persons, however, has not been determined.

In addition to improving muscle function, dietary NO_3_^−^ supplementation has also been found to reduce blood pressure in a number of subject groups (17). However, a recent meta-analysis concluded that dietary NO_3_^−^ does not reduce blood pressure in subjects ≥ 65 y of age (18). It was hypothesized that this may be due to variations in the oral microbiota and/or other factors (e.g., gastric acid production) that could influence the extent of reduction of NO_3_^−^ to NO_2_^−^(18). Alternatively, an age-related decline in the expression of the putative NO_3_^−^ transporter sialin could limit the ability of the salivary glands of older individuals to concentrate and resecrete NO_3_^−^ into the oral cavity (19). In either case, larger doses of NO_3_^−^ may be required to elicit significant increases in plasma NO_2_^−^ levels and NO bioavailability, and hence a decrease in blood pressure, in older persons. As with muscle function, however, the effect of NO_3_^−^ dose on changes in blood pressure in older individuals has not been tested.

The purpose of the present study was to determine the effects of varying doses of dietary NO_3_^−^ on changes in muscle speed and power and blood pressure in healthy older men and women. Based on previous research (20), we hypothesized that a moderate dose of NO_3_^−^ would increase muscle speed and power in such individuals, which would not be further enhanced at a higher dose of NO_3_^−^. Blood pressure, on the hand, would only be significantly reduced at a higher dose of NO_3_^−^.

## Methods

### Subjects

Nine healthy, community-dwelling older subjects (one man, eight women) with mean (±SE) age, height, weight, and body mass index of 70±1 y, 1.68±0.03 m, 68.7±4.2 kg, and 24.3±1.3 kg/m^2^ participated in this study. Potential subjects were assessed during an initial visit to the Clinical Research Center that included a health history and physical examination, resting electrocardiogram, phlebotomy for determination of screening/phenotyping laboratories (CBC, CMP, fasting insulin and lipids), and practicing the entire isokinetic dynamometry testing protocol (see below). Individuals were excluded if they met any of the following criteria: not between the ages of 65–79 y, unable to provide informed consent, were pregnant or lactating, had a history of major metabolic (Type I or II diabetes, thyroid disorders), neuromuscular (e.g., cervical spondylotic radiculomyelpathy, lumbar spondylosis, amyotrophic lateral sclerosis, Guillain-Barré syndrome, acquired demyelinating polyneuropathies), cardiovascular (e.g., moderate or severe valvular disease, myocardial infarction/ischemia, myocardial/pericardial disease, stage II or greater hypertension, heart failure), renal (eGFR < 60 mL/min/1.73 m^2^ or 61–90 mL/min/1.73m^2^ and albumin:creatine ratio > 30), or liver (i.e., SGOT/SGPT > 2x normal) disease, were anemic (hematocrit < 30%), or had any other contraindications to vigorous exercise. They were also excluded from the study if they currently smoked or were taking proton pump inhibitors, antacids, xanthine oxidase inhibitors, or were on hormone replacement therapy, as these can interfere with the reduction of NO_3_^−^ to NO_2_^−^ and then NO (21–23). Individuals on phosphodiesterase inhibitors were also excluded, as these can potentiate the effects of NO (24). The study was approved by the Human Subjects Office at Indiana University, and written, informed consent was obtained from each subject.

### Experimental design and protocol

Following completion of the initial screening visit, eligible volunteers were studied using a randomized, double-blind, placebo-controlled crossover design. They were asked to avoid sympathetic stimulation (caffeine, alcohol) and fast for 12 h prior to testing. Upon arrival at the University Hospital Clinical Research Center (CRC), blood pressure, heart rate, and the amount of NO in the breath were measured. An intravenous catheter was then placed, and a baseline blood sample was obtained for subsequent determination of plasma NO_3_^−^and NO_2_^−^ concentrations as described below. The subject then ingested 3.3 mL/kg of a commercial concentrated beetroot juice (BRJ) supplementation (Beet It Sport^®^, James White Drinks, Ipswich, UK) either: 1) essentially devoid of NO_3_^−^ (placebo); 2) containing (based on direct measurement) 121 µmol/mL NO_3_^−^ (thus total dose = 400 µmol/kg); or 3) an equal mixture of the placebo and NO_3_^−^ containing BRJ products (total dose = 200 µmol/kg). Hemodynamic data and breath/blood samples were collected again after 1 and 2 h of quiet rest, after which muscle contractile function was determined as described below. After a 10 min recovery period, heart rate and blood pressure were again measured and the final breath and blood samples were collected, then the subject was fed lunch and released from the CRC. They subsequently returned to the hospital and repeated the above procedures two more times receiving the other doses, with a 1–2 wk washout period between visits.

### Measurement of muscle contractile function

Maximal knee extensor power (Pmax, in W/kg) and velocity (Vmax, in rad/s) were measured using an isokinetic dynamometer (Biodex System 4 Pro^®^, Biodex Medical Systems, Shirley, NY) as previously described in detail (11). Briefly, the femoral condyle of the subject’s dominant leg was aligned with the axis of rotation of the dynamometer and their torso, waist, thigh, and lower leg tightly restrained with straps to prevent extraneous movement. The subject then performed maximal knee extensions at angular velocities spanning the ascending limb of the power-velocity relationship, i.e., at 0, 1.57, 3.14, 4.71, and 6.28 rad/s (0, 90, 180, 270, and 360 º/sec). Three maximal repetitions were executed at each velocity, with 2 min of rest allowed between each set of contractions. Subjects were instructed to attempt to move as quickly and as forcefully as possible during each repetition, and strong verbal encouragement was provided throughout the testing. To eliminate artifacts, the isokinetic dynamometer data were windowed, filtered, and smoothed as previously described (11), after which peak power at each velocity was calculated by multiplying the peak torque by velocity. Pmax and Vmax were then determined by fitting an inverted parabola to the peak power-velocity relationship (11).

### Measurement of breath NO and plasma NO_3_^−^, NO_2_^−^, and 3-NT

Breath NO was measured using a portable electrochemical analyzer (NIOX VERO^®^, Circassia Pharmaceuticals, Mooresville, NC). Plasma NO_3_^−^ and NO_2_^−^ were measured using a dedicated high performance liquid chromatography system (ENO-30, Eicom USA, San Diego, CA) as previously described in detail (25). Briefly, 25 µL of thawed plasma was mixed 1:1 with methanol, centrifuged at 4°C for 10 minutes at 10,000 g, and a 10 µL aliquot of the protein-poor supernatant injected into the HPLC. Plasma NO_3_^−^ and NO_2_^−^ concentrations were calculated based on standard curves generated using NIST-traceable standard solutions. Plasma 3-NT levels were determined using a commercial ELISA kit (OxiSelect^™^, Cell Biolabs, San Diego, CA).

### Sample size

The required sample sizes to detect changes in Pmax and Vmax (the primary outcome variables) were estimated based on the respective effect sizes observed in our recent study (16) and assuming α = 0.05 and 1-β = 0.90. These calculations indicated that n = 11 would be required to detect changes in Pmax, whereas n = 6 would be required to detect changes in Vmax. However, we were only able to study n = 9 subjects before all non-essential research at our institution was halted due to the COVID-19 pandemic.

### Statistical analyses

Statistical analyses were performed using GraphPad Prism version 8.4.1 (GraphPad Software, La Jolla, CA). Normality of data distribution was tested using the D’Agostino-Pearson omnibus test. Peak torque/power data and hemodynamic, breath NO, plasma NO_3_^−^, NO_2_^−^, and 3-NT data were analyzed using two-way (i.e., dose x velocity or dose x time, respectively) repeated measures ANOVA. Changes from baseline (average of 2 h and 10 min post-exercise values) in the latter variables as well as absolute values and changes in Vmax and Pmax were analyzed using one-way ANOVA. Post-hoc testing was performed using the Holm-Šidák multiple comparison procedure. P< 0.05 was considered significant.

## Results

### Markers of NO bioavailability

Plasma NO_3_^−^ and NO_2_^−^ concentrations and breath NO levels did not change following ingestion of the placebo BRJ, whereas ingestion of BRJ containing 200 µmol/kg of NO_3_^−^ resulted in marked elevations in all three (Table 1, Fig. 1A-C). The magnitude of the increases in plasma NO_3_^−^ and NO_2_^−^ over baseline were comparable to those observed in previous studies using similar doses of NO_3_^−^ (e.g., Ref. 20). Ingestion of BRJ containing twice as much NO_3_^−^ (i.e., 400 µmol/kg) resulted in almost exactly double the increase in plasma NO_3_^−^ and NO_2_^−^ concentration (Table 1, Fig. 1A and 1B). In contrast, the level of NO in breath increased less after the higher vs. the lower dose of NO_3_^−^ (Table 1, Fig. 1C), although it was still elevated compared to the placebo trial. Plasma 3-NT concentrations initially decreased then gradually increased during all three trials (P< 0.05), but these changes tended to be greater during the placebo and low dose trials versus the high dose trial (Table 1, Fig. 1D).

**Table 1.** Effect of varying doses of dietary NO_3_^−^ on plasma NO_3_^−^, plasma NO_2_^−^, breath NO, and plasma 3-nitrotyrosine in healthy older subjects.

|  | Dose of NO<sub>3</sub><sup>-</sup><br>(μmol/kg) | Pre <u>ingestion</u> | Time of measurement |  |  |
| --- | --- | --- | --- | --- | --- |
|  |  |  | 1 h post<br><u>ingestion</u> | 2 h post<br><u>ingestion</u> | 10 min post<br><u>exercise</u> |
| Plasma NO<sub>3</sub><sup>-</sup><br>(μmol/L) | 0 | 55±10 | 57±5 | 67±6 | 66±5 |
|  | 200 | 40±5 | 523±52<sup>‡</sup> | 624±53<sup>‡</sup> | 628±47<sup>‡</sup> |
|  | 400 | 34±5 | 793±73<sup>‡¶</sup> | 1184±89<sup>‡¶</sup> | 1296±94<sup>‡¶</sup> |
| Plasma NO<sub>2</sub><sup>-</sup><br>(μmol/L) | 0 | 0.31±0.05 | 0.30±0.02 | 0.33±0.04 | 0.35±0.05 |
|  | 200 | 0.31±0.04 | 0.46±0.07 | 0.53±0.07<sup>*</sup> | 0.82±0.18 |
|  | 400 | 0.34±0.06 | 0.48±0.06 | 0.95±0.18<sup>*§</sup> | 1.22±0.15<sup>*</sup> |
| Breath NO (ppb) | 0 | 19±3 | 22±3 | 25±3 | 21±3 |
|  | 200 | 18±3 | 39±6<sup>*</sup> | 43±7<sup>*</sup> | 50±10<sup>*</sup> |
|  | 400 | 18±3 | 31±4<sup>†</sup> | 36±4<sup>†</sup> | 36±6<sup>*</sup> |
| 3-Nitrotyrosine<br>(nmol/L) | 0 | 177±20 | 153±18 | 170±19 | 217±27 |
|  | 200 | 170±22 | 158±19 | 176±23 | 216±27 |
|  | 400 | 237±25 | 209±22 | 198±26 | 250±34 |

Values are mean±SE for n=9. Significantly different from 0 μmol/kg dose: <sup>\*</sup>P<0.05, <sup>†</sup>P<0.01,
<sup>‡</sup>P<0.001. Significantly different from 200 μmol/kg dose: <sup>§</sup>P<0.05, <sup>¶</sup>P<0.001.

**Figure 1.**
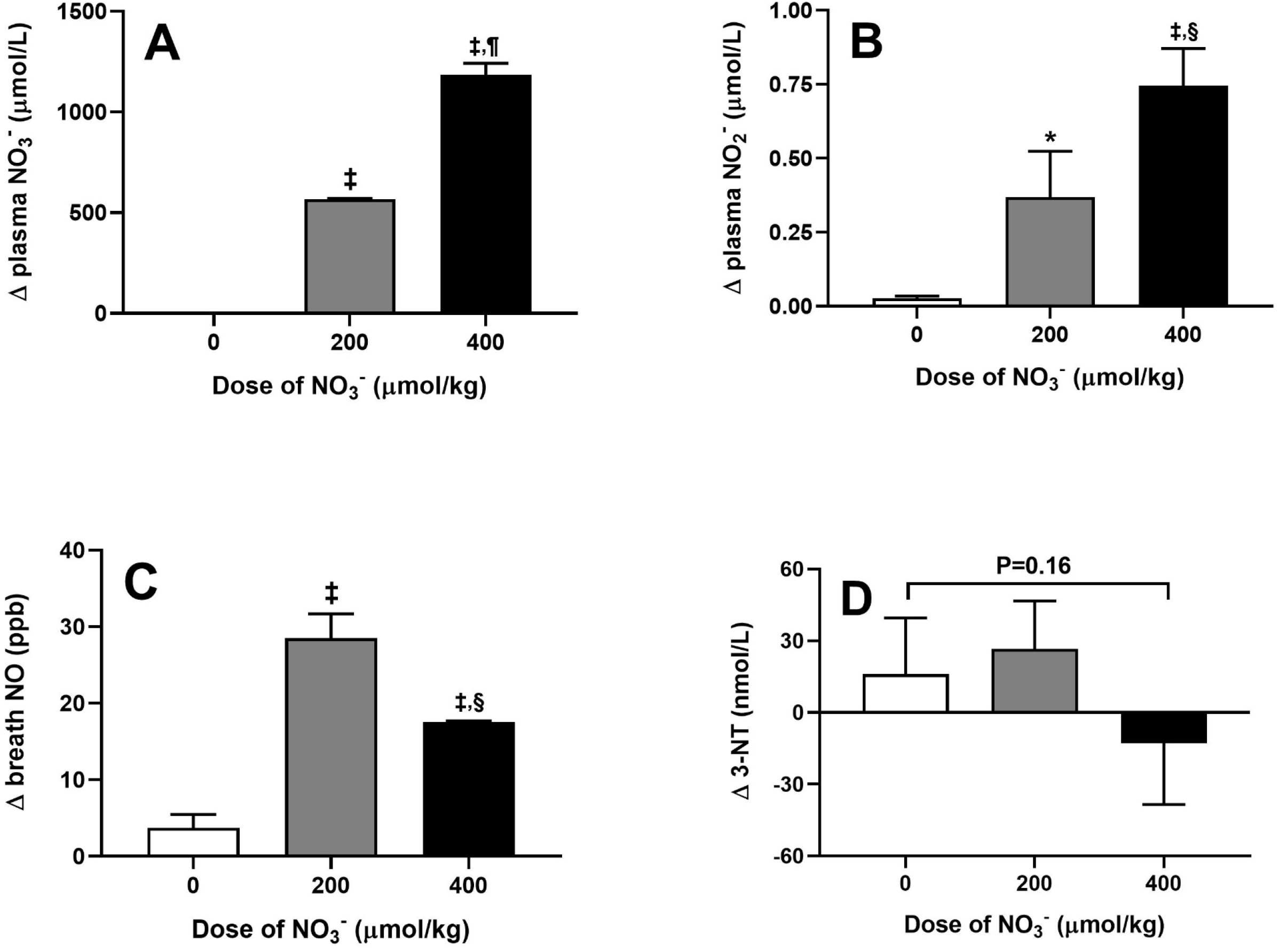
Changes (Δ) in plasma nitrate (NO_3_^−^; *panel A*), nitrite (NO_2_^−^; *panel B*), breath nitric oxide (NO; *panel C*), and plasma 3-nitrotyrosine (3-NT; *panel D*) in response to varying doses of dietary NO_3_^−^ in healthy older subjects. Values are mean±SE for n = 9. Significantly different from 0 µmol/kg dose:^*^P< 0.05,^‡^P< 0.001. Significantly different from 200 µmol/kg dose:^§^P< 0.05, ^¶^P< 0.001.

### Muscle contractile function

There were no significant effects of NO_3_^−^ ingestion on knee extensor peak torque, and hence peak power, at low to moderate angular velocities (i.e., at 0, 1.57, 3.14, or 4.71 rad/s; Table 2). In keeping with our previous results (16), however, ingestion of BRJ containing a moderate amount of NO_3_^−^ (i.e., 200 µmol/kg) resulted in a significant increase in peak torque, and hence peak power, at the highest velocity tested (i.e., 6.28 rad/s) (Table 2). This beneficial effect of acute NO_3_^−^ supplementation on muscle contractile function was lost when the NO_3_^−^ dose was doubled to 400 µmol/kg (Table 2). Correspondingly, Pmax (in W/kg) as calculated from these data was significantly higher (P< 0.05) after the low dose (3.9±0.4) compared to the placebo (3.7±0.4) or high dose (3.7±0.4). Vmax (in rad/s) also tended higher (P = 0.08) in the low (11.9±0.7) compared to the placebo (10.8±0.8) or high (11.2±0.8) trials. Eight out of nine subjects (including the one man) achieved a higher Pmax and Vmax after ingestion of the lower vs. the higher dose of NO_3_^−^ (Figs. 2A-D).

**Table 2.** Effect of varying doses of dietary NO_3_^−^ on peak knee extensor torque and power as a function of velocity in healthy older subjects.

|  |  | Angular velocity (rad/s) |  |  |  |  |
| --- | --- | --- | --- | --- | --- | --- |
| Dose of NO<sub>3</sub><sup>-</sup> |  |  |  |  |  |  |
| <u>(μmol/kg)</u> |  | <u>0</u> | <u>1.57</u> | <u>3.14</u> | <u>4.71</u> | <u>6.28</u> |
| Peak torque<br>(Nm/kg) | 0 | 1.79±0.15 | 1.21±0.10 | 0.91±0.06 | 0.75±0.06 | 0.55±0.06 |
|  | 200 | 1.86±0.15 | 1.24±0.07 | 0.92±0.06 | 0.78±0.06 | 0.61±0.06<sup>†</sup> |
|  | 400 | 1.78±0.12 | 1.24±0.07 | 0.90±0.05 | 0.74±0.06 | 0.56±0.06 |
| Peak power<br>(W/kg) | 0 | 0.00±0.00 | 1.97±0.15 | 2.91±0.22 | 3.59±0.30 | 3.46±0.45 |
|  | 200 | 0.00±0.00 | 1.95±0.11 | 2.89±0.20 | 3.69±0.27 | 3.83±0.37<sup>†</sup> |
|  | 400 | 0.00±0.00 | 1.94±0.11 | 2.84±0.17 | 3.48±0.28 | 3.49±0.39 |

Values are mean±SE for n=9. Significantly different from 0 or 400 μmol/kg doses: <sup>†</sup>P<0.01.

**Table 3.** Effect of varying doses of dietary NO_3_^−^ on heart rate and blood pressure in healthy older subjects.

| | Dose of NO<sub>3</sub><sup>-</sup><br>(μmol/kg) | Time of measurement | | | |
| --- | --- | --- | --- | --- | --- |
|  |  | Pre <u>ingestion</u> | 1 h post<br><u>ingestion</u> | 2 h post<br><u>ingestion</u> | 10 min post<br><u>exercise</u> |
| Heart rate<br>(beats/min) | 0 | 73±4 | 72±3 | 70±3 | 73±3 |
| | 200 | 71±4 | 74±3 | 70±3 | 73±2 |
| | 400 | 72±3 | 70±3 | 71±3 | 74±3 |
| Systolic blood<br>pressure<br>(mmHg) | 0 | 131±5 | 122±4 | 122±3 | 125±4 |
| | 200 | 130±4 | 122±6 | 121±5 | 119±6 |
| | 400 | 134±4 | 121±4 | 118±5 | 118±6 |
| Diastolic blood<br>pressure<br>(mmHg) | 0 | 70±2 | 68±2 | 68±3 | 69±2 |
| | 200 | 69±4 | 67±3 | 67±3 | 68±4 |
| | 400 | 69±3 | 62±3 | 64±4 | 65±3 |
| Mean arterial<br>pressure<br>(mmHg) | 0 | 90±2 | 86±2 | 86±2 | 87±2 |
| | 200 | 90±3 | 85±3 | 85±3 | 85±3 |
| | 400 | 90±3 | 82±3 | 82±3 | 82±3 |

Values are mean ± SE for n=9.

**Figure 2.**
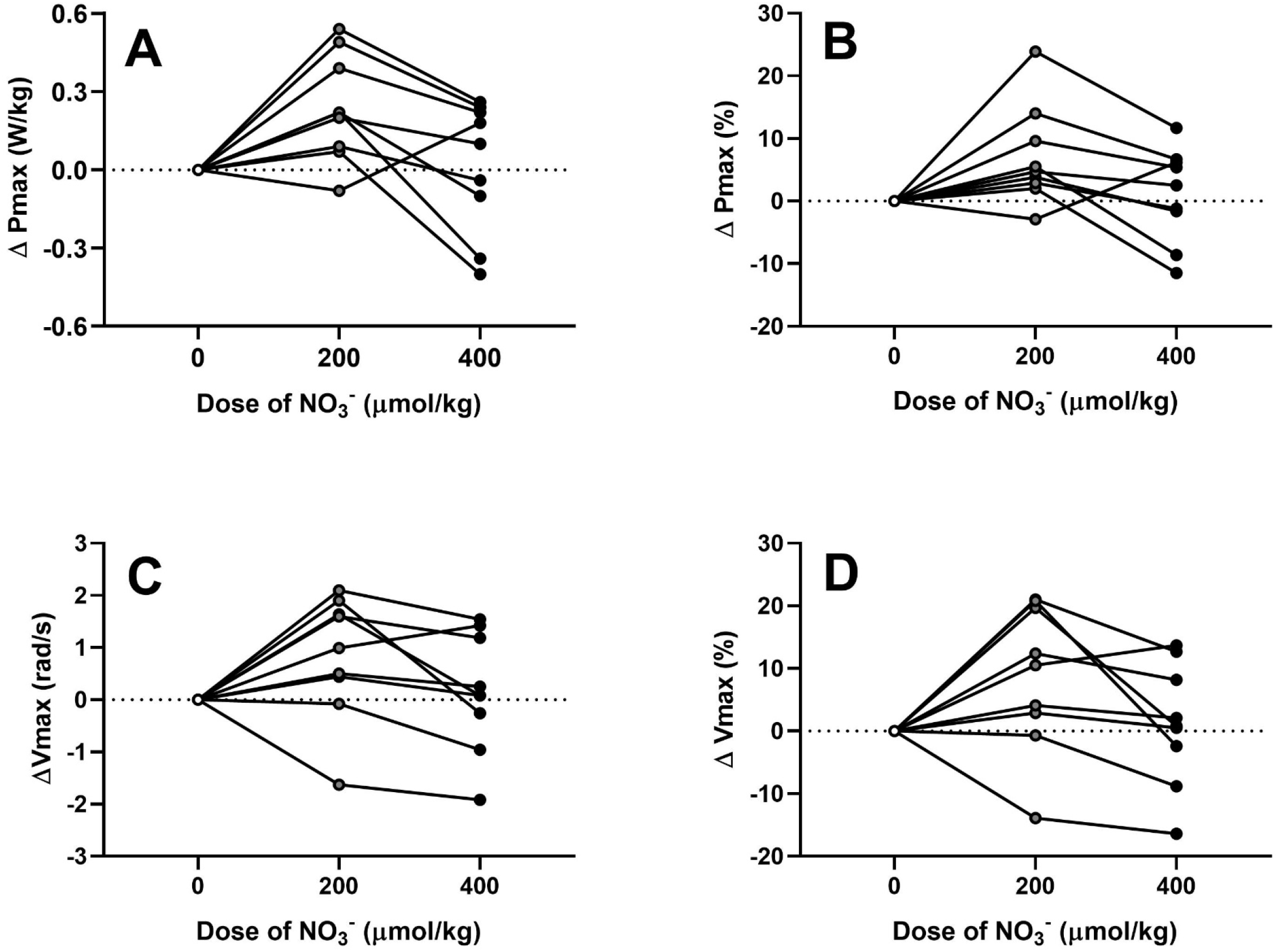
Individual absolute and relative changes (Δ) in maximal knee extensor power (Pmax; *panels A and B*) and velocity (Vmax; *panels C and D*) in response to varying doses of dietary nitrate (NO_3_^−^) in healthy older subjects.

### Hemodynamics

Heart rate did not change significantly over time or in response to ingestion of varying amounts of NO_3_^−^ (Table 3). Systolic and mean blood pressure, on the other hand, decreased significantly over time (both P< 0.01 for main effect), with such changes tending to be greatest in the high dose trial (Table 3). However, neither the treatment main effects nor the treatment x time interaction effects were statistically significant. Diastolic blood pressure did not change significantly with either time or treatment.

## Discussion

The purpose of the present study was to determine the effect of varying doses of dietary NO_3_^−^ on changes in muscle contractile function and blood pressure in healthy older individuals. We found that a moderate dose of NO_3_^−^ (i.e., 200 µmol/kg) resulted in measurable improvements in maximal knee extensor speed and power, but no significant changes in blood pressure. A larger dose of NO_3_^−^ (i.e., 400 µmol/kg) also did not significantly alter blood pressure, whereas muscle speed and power actually regressed towards, and in some individuals even below, baseline values. This is the first dose-response study of the effects of acute dietary NO_3_^−^ supplementation on muscle contractile function in any population, and the first to specifically examine changes in blood pressure in older subjects.

We have previously demonstrated that dietary NO_3_^−^ supplementation can enhance muscle contractility in a variety of subject populations (11–14). This includes healthy elderly men and women, in whom acute intake of 13.4±0.5 mmol (182±10 µmol/kg) of NO_3_^−^ increased maximal knee extensor speed and power by 11±2 and 4±4%, respectively (16). The results of the present study are similar, in that Vmax and Pmax improved by 8±4 and 7±3%, respectively, after ingestion of BRJ containing 200 µmol/kg of NO_3_^−^. The somewhat greater increase in Pmax in the present investigation vs. our previous study could be due to the slightly larger dose of NO_3_^−^provided. Indeed, in our previous study we observed that individuals who received less than ∼150 µmol/kg of NO_3_^−^ were less likely to show significant improvements in muscle function (unpublished observations). Alternatively, it could be due to the greater percentage of women in the present study (i.e., 89 vs. 50%), as we have found that women tend to be more likely to benefit from NO_3_^−^ intake (12). Regardless, as previously discussed these improvements in muscle function are substantial relative to their normal rate of decline with aging, and in fact are roughly comparable to those resulting from several months of traditional resistance exercise training (cf. Ref. 26 for review). As such, dietary NO_3_^−^ supplementation could potentially have a measurable impact on the functional capacity (e.g., walking speed, chair rise time, stair climbing ability) of older persons during daily life, thus minimizing the risk of disability and dependence.

Given the latter, it is imperative to determine the optimal dose (and timing and source, i.e., BRJ vs. a salt) of NO_3_^−^ for enhancing muscle contractile function in older persons. Previous dose-response studies in younger untrained or moderately-trained individuals have found that providing 6–10 mmol (roughly 100–150 µmol/kg) of NO_3_^−^ is generally sufficient to improve economy and/or performance during endurance exercise (20,27–29), with no further improvements at higher doses (i.e., up to 16.8 mmol, or ∼225 µmol/kg) (20). As implied above, however, it appears that somewhat higher doses of NO_3_^−^ may be required to increase muscle speed and power, at least in older men and women. Thus, in addition to studying a NO_3_^−^ dose of 200 µmol/kg, we also tested subjects after they had consumed double this dose, i.e., 400 µmol/kg. Based on our previous research and that of Wylie et al. (20), we anticipated that this would result in a quantitatively-similar improvement in muscle contractile function, i.e., that both 200 and 400 µmol/kg would fall on a plateau in the dose-response relationship. Surprisingly, however, both Vmax and Pmax actually regressed at the higher dose, such that on average they were not significantly different from the placebo trial. This pattern was highly consistent across subjects, with all but one of the subjects performing best with the lower dose, and several of the subjects actually performing worse following the higher dose than during the placebo trial. To our knowledge, this is the first study to demonstrate a possible impairment in exercise performance following ingestion of dietary NO_3_^−^, although both Larsen et al. (30) and Bescós et al. (31) have previously reported that NO_3_^−^ ingestion can reduce VO_2_max.

The mechanism responsible for the above unanticipated finding is not clear. It does not, however, appear to be due to failure of the higher dose of NO_3_^−^ to further augment plasma NO_3_^−^and NO_2_^−^ levels, as both rose almost exactly twice as much following the 400 vs. the 200 µmol/kg dose. This is consistent with the essentially complete bioavailability of NO_3_^−^ from ingested vegetables (32). Interestingly, however, the availability of NO itself, at least as measured in the breath, rose less after the higher vs. the lower dose, and thus was significantly lower in the former vs. the latter experiments. We therefore measured plasma 3-NT concentrations, as another marker of the rate of NO production but also to determine whether the higher dose of NO_3_^−^ (which averaged 28.7±2.7 mmol) resulted in “peaking over” of muscle contractile function due to an increase in nitrosative stress. This possibility was suggested by the recent results of Gholami et al. (33), who reported that ingestion of 24, but not 8 or 16, mmol of NO_3_^−^ resulted in a dramatic increase in plasma peroxynitrite levels in well-trained subjects following a maximal incremental exercise test. In the present study, however, the pattern of change in plasma 3-NT instead generally resembled that of breath NO, i.e., tending to increase slightly after the lower dose but decrease slightly after the higher dose of NO_3_^−^.

We interpret the above findings as follows: the lower dose of NO_3_^−^ significantly increased plasma NO_3_^−^ concentrations, which led to a rise in plasma (and presumably tissue (34)) NO_2_^−^levels, and hence an increase in NO bioavailability. As a result of the latter, both muscle contractile function (i.e., Vmax and Pmax) and plasma 3-NT concentrations were elevated (the latter insignificantly) compared to the placebo trial. Conversely, the higher dose of NO_3_^−^ resulted in even greater increases in plasma NO_3_^−^ and NO_2_^−^ concentrations, but NO itself actually increased less than after the lower dose, ostensibly as a result of NO_3_^−^ outcompeting NO_2_^−^ for reduction by XOR. Consequently, muscle speed and power did not change significantly, whereas plasma 3-NT concentrations actually tended to decline. This scenario, which is shown schematically in Figure 3, is partially based on the observations of Damacena-Angelis et al. (35), who found that higher concentrations of NO_3_^−^ can inhibit XOR-mediated reduction of NO_2_^−^ to NO, and thus partially attenuate the blood pressure lowering effect of NO_2_^−^ in rats. Although speculative, the above hypothesis could explain the present observations. Alternatively, it is possible that the high concentrations of NO_3_^−^ and/or NO_2_^−^ present after ingestion of the higher dose led to a direct inhibitory effect on muscle contractile function, e.g., by enhancing transnitrosylation of myosin (36). This would not, however, account for the lower NO levels observed following ingestion of the higher dose of NO_3_^−^.

**Figure 3.**
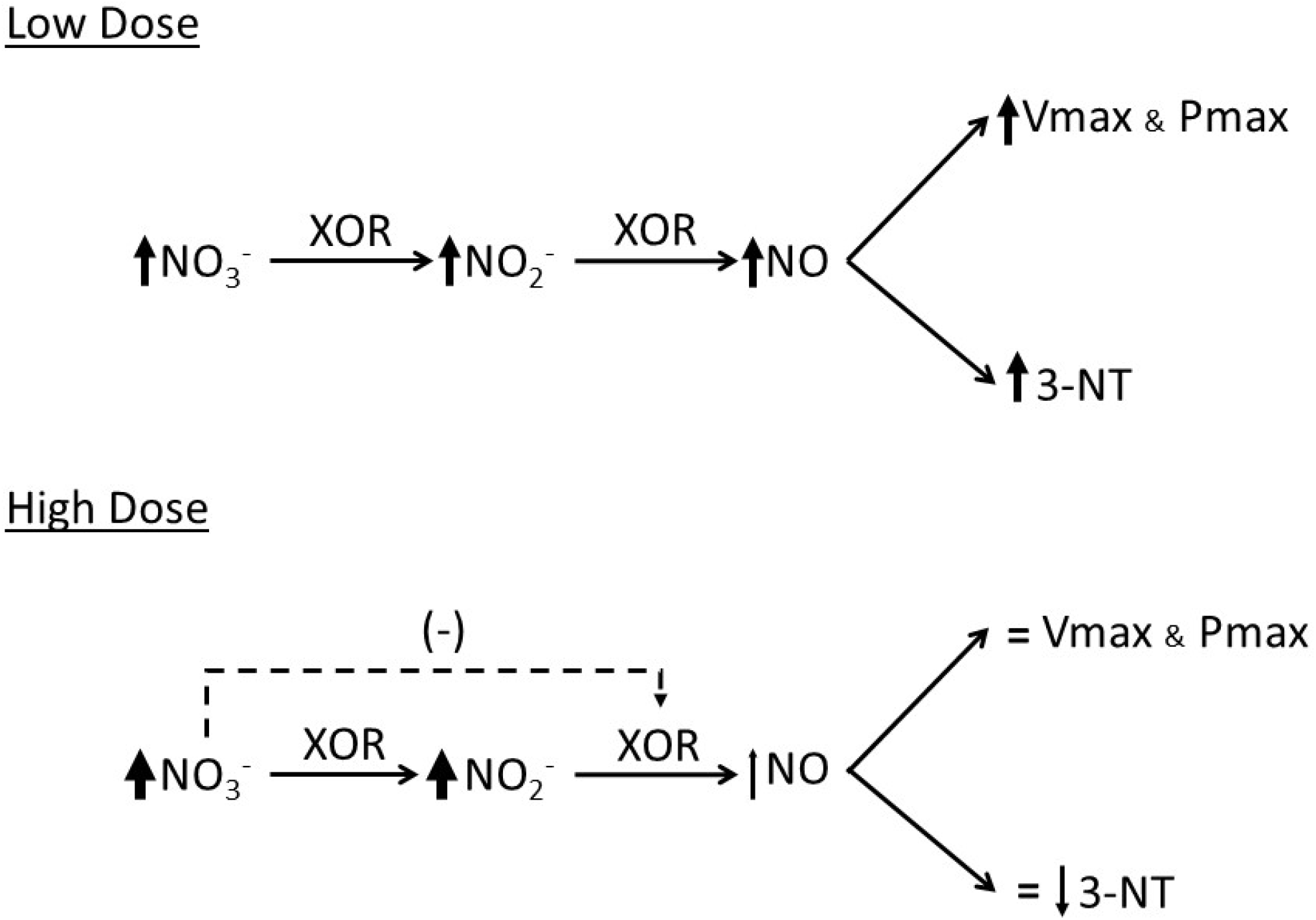
Proposed explanation for the effect of varying doses of dietary nitrate (NO_3_^−^) on muscle contractile function in healthy older subjects. A lower dose of NO_3_^−^ (i.e., 200 µmol/kg) is hypothesized to result in a significant increase in muscle NO_3_^−^ concentration, which is reduced by xanthine oxidoreductase (XOR) to nitrite (NO_2_^−^) and then to nitric oxide (NO), leading to significant improvements in maximal knee extensor power (Pmax) and velocity (Vmax) and also to an increase in 3-nitrotyrosine (3-NT) formation. In contrast, a higher dose of NO_3_^−^ (i.e., 400 µmol/kg) actually results in less of an increase in NO production, due to substrate competition between NO_3_^−^ and NO_2_^−^ for XOR. As a result, Pmax, Vmax, and 3-NT are unchanged, or possibly even diminished.

In contrast to the improvements in muscle speed and power observed following ingestion of the lower dose of NO_3_^−^, blood pressure was unaltered by either the lower or the higher dose. This finding is consistent with a recent meta-analysis by Siervo et al. (18) but runs counter to the results of other studies of older subjects (37–39). The reason for these divergent results is uncertain. The amount of NO_3_^−^ provided in the present experiments is much higher than used in these previous studies, such that an inadequate dose can seemingly be ruled out. It is possible that the subjects we studied were resistant to the potential blood pressure-lowering effects of dietary NO_3_^−^ due to excessive oxidative stress, since NO_3_^−^ plus ascorbic acid but not NO_3_^−^ alone has been reported to reduce systolic and mean arterial blood pressure in older adults (40). It is also possible, however, that the failure to detect a statistically-significant NO_3_^−^-induced reduction in blood pressure in the present study is simply a type 2 statistical error, due to the well-known variability of blood pressure measurements combined with a relatively small number of subjects. This is so because 1) changes in blood pressure from baseline were numerically greater after NO_3_^−^ ingestion (i.e., −8±3, −11±4, and −16±4, −1±2, −2±2, and −4±2, and −4±2, −5±2, and −8±2 mmHg for systolic, diastolic, and mean arterial pressure in the placebo, 200 µmol/kg, and 400 µmol/kg trials, respectively), and 2) post-hoc analysis of the data demonstrated that, given the observed effect sizes, the achieved powers (i.e., 1-β) with n = 9 subjects were only 0.64, 0.67, and 0.83, respectively.

There are both strengths and weaknesses to the present study. A major strength is the use of a highly-reliable method for assessing maximal muscle contractile properties, even in older subjects. For example, as calculated from the ANOVA the intraclass correlation coefficient (ICC) for Pmax was 0.98. This helps explain why we were able to detect statistically-significant changes in muscle contractile function but not in, e.g., systolic blood pressure (ICC = 0.70). Another strength is that, unlike in previous dose-response studies (20,27–29), in the present experiments the dose of NO_3_^−^ was adjusted relative to body mass, thus at least theoretically minimizing inter-subject variability in peak plasma NO_3_^−^ concentrations due to differences in the volume of distribution. At the same time, we kept the volume of BRJ the same across trials, thus eliminating differences in the amount of other potentially biologically-active compounds (e.g., carotenoids, betalains, bioflavonoids, ascorbic acid) as a source of variability. As discussed above, a weakness of our study is that it was slightly underpowered to detect possible differences in blood pressure due to NO_3_^−^ ingestion. Another weakness is that by studying only two doses of NO_3_^−^, i.e., 200 and 400 µmol/kg, it is not possible to determine whether the optimal dose is somewhat less than 200 µmol/kg or somewhere between 200 and 400 µmol/kg. At a minimum, however, the present data provide a starting point for further experiments.

In summary, in the present study we have examined the dose-response relationship between dietary NO_3_^−^ supplementation and changes in both muscle contractile function and blood pressure in healthy older subjects. We found that although blood pressure was unchanged, ingestion of a lower, but not a higher, dose of NO_3_^−^ significantly increased maximal knee extensor power, with these divergent results seemingly reflecting the failure of the higher dose to sufficiently elevate NO bioavailability. Future studies should use the tools and approaches commonly employed in pharmaceutical development (e.g., pharmacokinetic-pharmacodynamic modeling) to further refine this possible treatment for muscle dysfunction in older persons.

## Data Availability

Data available upon request from corresponding author.

